# Early-life hallmarks of polygenic liability to adult internalizing-cardiometabolic multimorbidity

**DOI:** 10.64898/2026.05.01.26352146

**Authors:** Ruby S. M. Tsang, Daniel Stow, Ioanna K. Katzourou, LINC Consortium, Marianne B. M. van den Bree, Golam M. Khandaker, Nicholas J. Timpson

**Affiliations:** MRC Integrative Epidemiology Unit, University of Bristol, Bristol, UK; Population Health Sciences, Bristol Medical School, University of Bristol, Bristol, UK; Centre for Academic Mental Health, Population Health Sciences, University of Bristol, UK; Wolfson Institute of Population Health, Queen Mary University of London, London, UK; Centre for Neuropsychiatric Genetics and Genomics, Division of Psychological Medicine and Clinical Neurosciences, Cardiff University, Cardiff, UK; Wales Kidney Research Unit, Cardiff University, Cardiff, UK; Neuroscience and Mental Health Innovation Institute, Division of Psychological Medicine and Clinical Neurosciences, Cardiff University, Cardiff, UK; NIHR Bristol Biomedical Research Centre and NIHR Bristol Clinical Research Facility, University Hospitals Bristol and Weston NHS Foundation Trust, Bristol, UK; Avon and Wiltshire Mental Health Partnership NHS Trust, Bristol, United Kingdom

**Author notes:** Corresponding author: Ruby Tsang. A list of consortium members and their affiliations is included at the end of the paper (LINC Consortium).

**Keywords:** multimorbidity, mental health, cardiometabolic disease, polygenic risk score, inflammation, ALSPAC

## Abstract

**Background:** Internalizing disorders and cardiometabolic disease are common conditions that frequently co-occur in later life and may be attributed to shared genetic influences. While phenotypic effects of polygenic liability of adult disorders may emerge early in life, studies have not investigated this in the context of multimorbidity. This study set out to investigate early manifestations of polygenic liability to adult internalizing-cardiometabolic multimorbidity (ICM-MM) in a UK population birth cohort.

**Methods:** We used data from 5,934 individuals in the Avon Longitudinal Study of Parents and Children (ALSPAC). We modelled trajectories of 12 mental and cardiometabolic health outcomes using mixed effects models, and investigated effects of adult ICM-MM polygenic liability on these trajectories. We also investigated associations of adult ICM-MM polygenic liability with circulating inflammatory proteins (Olink Target 96 Inflammation panel) at ages 9 and 24.

**Results:** Adult ICM-MM polygenic liability is associated with cardiometabolic traits and inflammation, and with changes in depressive symptoms and cardiometabolic traits over time in childhood through to early adulthood. A notable early life biological footprint is inflammation. We found that higher ICM-MM polygenic liability is consistently associated with higher interleukin-6 (IL6), tumor necrosis family superfamily member 14 (TNFSF14) and hepatocyte growth factor (HGF) levels in both childhood and early adulthood.

**Conclusions:** Adult ICM-MM polygenic liability manifests early in life through changes in mental and cardiometabolic health and blood biomarkers, especially in increases of circulating inflammatory proteins related to obesity, immune cell chemotaxis and migration that may contribute to disease pathogenesis by seeding inflammation in relevant tissues.

## Introduction

Multimorbidity is defined as the co-occurrence or lifetime occurrence of two or more chronic conditions, which could be either a physical non-communicable disease, a mental health condition, or an infectious disease (1, 2). It is estimated that approximately 40% of the general population have multimorbidity (3, 4), and the number of individuals with multimorbidity are projected to almost double by 2050 (5, 6). The rapidly increasing burden of multimorbidity poses significant public health challenges, as multimorbidity is associated with higher healthcare utilization and costs, functional impairment, poorer health-related quality of life and premature mortality (7).

Internalizing disorders (e.g., depression and anxiety) and cardiometabolic disease (e.g., coronary heart disease, stroke and type 2 diabetes) are consistently among the leading causes of health-related disability worldwide (8). These conditions frequently co-occur and are bidirectionally associated (9). Depression and anxiety have been shown to be associated with increased risk of coronary heart disease (10, 11), stroke (12, 13), type 2 diabetes (14, 15), and vice versa (15–18). As these conditions have a heritable component (19–22), common co-occurrence may in part be due to shared genetic influences across the life course. Twin studies have reported modest genetic correlations of depression with body mass index (BMI) (23), coronary artery disease (24) and type 2 diabetes (25), and these findings have also been supported by polygenic risk score (PRS) and genetic correlation analyses in a more recent study involving a large multi-cohort sample (26).

Many chronic conditions have origins in early life (27). However, the absence of deep longitudinal data spanning decades limits the possibility of conducting life course analyses to identify early manifestations and trajectories of chronic disease with a typical later-life onset. An individual’s susceptibility to internalizing disorders and cardiometabolic conditions can be partly captured by PRSs. Recent studies have shown that phenotypic effects of polygenic risk emerge early in life, with PRSs capable of accurately stratifying individuals across a range of cardiometabolic risk factors and biomarkers in childhood (28, 29). However, whether the same is true for adult internalizing-cardiometabolic multimorbidity (ICM-MM) is unknown. Given that depression and cardiovascular risk factors are among the most consistently reported risk factors associated with incident multimorbidity in adulthood (30), it is of interest to investigate whether polygenic liability to adult ICM-MM is associated with differences in, or accelerated development of, these risk factors earlier in life. Prospective longitudinal population-based cohorts represent a unique framework for studying early manifestations of genetic risk of later-onset diseases using a life course epidemiological approach.

The aim of this study is to leverage deep longitudinal data in the Avon Longitudinal Study of Parents and Children (ALSPAC), a United Kingdom (UK)-based birth cohort with over twenty years of follow-up, to investigate early manifestations of polygenic liability to adult ICM-MM. Here we examine the effects of a PRS of adult ICM-MM on trajectories of 12 selected mental and cardiometabolic health phenotypes (depressive symptoms, body mass index [BMI], fat mass index [FMI], lean mass index [LMI], systolic and diastolic blood pressure [SBP and DBP], high-density lipoprotein [HDL], non-HDL, triglycerides [TG], fasting glucose and insulin [FG and FI], and C-reactive protein [CRP]) in childhood to early adulthood, and associations with circulating inflammatory proteins.

## Methods and Materials

### Cohort

ALSPAC is a population-based prospective birth cohort (31–34). Pregnant women resident in Avon, UK with expected dates of delivery between April 1991 and December 1992 were invited to participate. The initial number of pregnancies enrolled was 14,541, of which 13,988 infants were alive at one year of age. Further recruitment took place when the oldest children were approximately seven years of age; the total sample size for analyses using any data collected after the age of seven is 15,447 pregnancies, of which 14,901 were alive at one year of age.

We included in the study sample unrelated individuals from Generation 1 of ALSPAC by applying a KING kinship coefficient cutoff of 0.0884, who are of European ancestry, and have both genotyping data and data on at least one outcome of interest available.

Please note that the study website contains details of all the available data through a fully searchable data dictionary and variable search tool: https://www.bristol.ac.uk/alspac/researchers/our-data/

### Exposure

#### Polygenic risk score for internalizing-cardiometabolic multimorbidity

The Lifespan Multimorbidity Research Collaborative (LINC) (https://www.cardiff.ac.uk/lifespan-multimorbidity-research-collaborative) defines ICM-MM as having a diagnosis of at least one of depression, anxiety or somatoform disorder *and* a diagnosis of at least one of obesity, hypertension, dyslipidemia, type 2 diabetes mellitus or chronic kidney disease. This definition is the result of discussion with primary and secondary care doctors, patient and public involvement advisors, as well as the broader research team. The conditions included are restricted to those that generally occur earlier in life than established cardiovascular disease, which will allow us to characterize the development of ICM-MM across the life course and identify opportunities for prevention and intervention.

In an earlier study, we tested associations of single-trait PRSs for the aforementioned conditions of interest and trained an ICM-MM PRS in UK Biobank (35). In brief, in that study we obtained summary statistics from the largest genome-wide association studies not including UK Biobank in their samples (a list of the GWASs used is in **Table S1**), and computed PRSs using PRS-CS (36) and the European ancestry 1000 Genomes external linkage disequilibrium reference panel. PRS-CS uses a Bayesian algorithm to infer posterior single nucleotide polymorphism (SNP) effect sizes under continuous shrinkage priors. The ICM-MM PRS was trained using elastic net regression, implemented within PRSmix (37) (a list of retained PRSs and their respective weights are in **Table S2**).

DNA samples from Generation 1 of ALSPAC were genotyped on the Illumina HumanHap550 quad chip, with imputation using the Haplotype Reference Consortium (HRC) panel. We computed single-trait PRSs in ALSPAC by first identifying common SNPs in UK Biobank and ALSPAC, then excluding mismatched and palindromic SNPs, and applying the SNP effect sizes from PRS-CS from our earlier study. We computed the ICM-MM PRS in ALSPAC by computing a weighted sum of the single-trait PRSs using the weights obtained from PRSmix. The ICM-MM PRS was used as the exposure in the main analysis of this study.

### Outcomes

We included 12 mental, cardiometabolic and inflammatory phenotypes as outcomes, selected based on literature on multimorbidity risk factors and availability of repeated measures data within ALSPAC. These comprised depressive symptoms (assessed using the Short Mood and Feelings Questionnaire [SMFQ]), BMI, FMI, LMI, SBP and DBP, HDL, non-HDL, TG, FG and FI, and CRP. Details of these phenotypes are outlined in **Supplementary Methods** and **Table S3**.

### Main longitudinal analysis

The ICM-MM PRS was standardized prior to analysis. All outcomes were natural log- or log1p-transformed prior to analysis so estimates reflect percentage change per one standard deviation increase in ICM-MM PRS. Age was calculated by dividing the age in months variable by 12 and used as the time variable in all models.

We first fitted mixed effects models with natural cubic splines using maximum likelihood estimation to estimate health trajectories over time. We included fixed effects of natural cubic splines of age and sex, and random intercepts (38). These models estimate the mean trajectory of each outcome while allowing for non-linear change over time and accounting for non-independence of repeated measurements within individuals, using all data from all eligible individuals under a missing at random (MAR) assumption. To facilitate interpretability of estimates, age was centered on the closest whole year of age at the earliest occasion of data collection for each outcome (i.e. seven for BMI, blood pressure and lipids; nine for FMI, LMI and CRP; 10 for depressive symptoms; and 15 for FG and FI). We tested models from two up to the lower of five or (*k*-1) degrees of freedom (*k* is the number of repeated measures available) and compared model fit using the Bayesian information criterion (BIC). Where a simpler model has a BIC less than 10 higher than the best-fitting model, the simpler model was selected. Knots were placed at equal quantiles of age. We then examined the effects of PRSs by including a PRS main effect and an age x PRS interaction effect in the models, adjusting for first 10 genetic principal components to account for potential population stratification. We controlled the false discovery rate (FDR) at 5% using the Benjamini-Hochberg procedure to account for multiple comparisons. For easier interpretation, we predicted values on the outcomes of interest for the 10^th^, 50^th^ and 90^th^ centiles, and then plotted the trajectories in their original scale.

### Sensitivity analysis

As a comparison built around existing PRSs designed to be used as integrated health predictors (related to our definition of ICM-MM), we conducted sensitivity analyses by running the same models with 1) another ICM-MM PRS trained using the same methods but with Genomics plc’s Enhanced Set of PRSs (39) as input (ICM-MM PRS_GPLC_; a list of retained PRSs and their respective weights are in **Table S4**, correlations with single-trait and ICM-MM PRS are in **Figure S1**), 2) all component single-trait PRSs, 3) a negative control with age-related macular degeneration PRS_GPLC_ (which showed no correlation with either ICM-MM PRS or ICM-MM PRS_GPLC_), and 4) by excluding CRP values >10mg/L as values over this threshold may indicate acute infections or injury in a subset of individuals (40).

### Secondary analysis

To more deeply characterize the biological profiles of individuals with high polygenic liability of ICM-MM, we conducted a further analysis to examine associations of the ICM-MM PRS with circulating inflammation and immune proteins in childhood and in early adulthood.

Heparin-stored plasma samples collected from the age 9 (non-fasting) and age 24 (fasting) research clinics were analyzed using the Olink Target 96 Inflammation panel (Olink Analysis Service, Uppsala, Sweden). This panel measures a selection of 92 proteins involved in inflammatory and immune response processes. Data were returned in normalized protein expression (NPX) values, Olink’s arbitrary unit on log_2_ scale. Samples that did not pass Olink’s quality control were removed but values below the limit of detection were kept in the analyses. Further details on pre-processing and quality control procedures applied in ALSPAC have been described elsewhere (41).

NPX values were rank-based inverse normal transformed prior to analysis. We tested associations between ICM-MM PRSs and 92 circulating inflammatory proteins using linear regressions, adjusting for age, sex and first 10 genetic principal components. We did not adjust for batch as there was no evidence of batch effects (41). We controlled the false discovery rate (FDR) at 5% using the Benjamini-Hochberg procedure to account for multiple comparisons. Proteins nominally associated with ICM-MM PRS were entered into the STRING database (https://www.string-db.org/; version 12.5) for visualization of protein-protein interactions and pathway analysis. We also conducted sensitivity analyses using the component single-trait PRSs.

### Software and packages

Extraction of phenotypic data was performed in Stata MP 18.5 (42). Genetic data were extracted using QCTOOL 2.2.0 and single-trait PRSs of LINC conditions were computed using PLINK 2.0. All statistical analyses were conducted using R 4.4.2 (43) in RStudio 2026.09.0 (44), with packages tidyverse (2.0.0), haven (2.5.5), lme4 (1.1-37), lmerTest (3.1-3), splines (4.4.2), broom.mixed (0.2.9.6) and broom (1.0.9). Plots were generated using corrplot (0.95), ggeffects (2.3.1), ggplot2 (3.5.2), ggpubr (0.6.1), ggrepel (0.9.6) and ggstats (0.10.0).

### Ethics

Ethical approval for the study was obtained from the ALSPAC Ethics and Law Committee and the Local Research Ethics Committees. Informed consent for the use of all data collected was obtained from participants following the recommendations of the ALSPAC Ethics and Law Committee at the time. Participants can contact the study team at any time to retrospectively withdraw consent for their data to be used. Study participation is voluntary and during all data collection sweeps, information was provided on the intended use of data. Further details are in **Supplementary Methods**.

## Results

### Sample characteristics

We included 5,934 individuals in this study, with sample sizes for the mixed effects models ranging from 3,493 to 5,800 (see study sample selection flow diagram in **Figure 1**). Selected characteristics of the included individuals are presented in **Table 1**.

**Figure 1.**
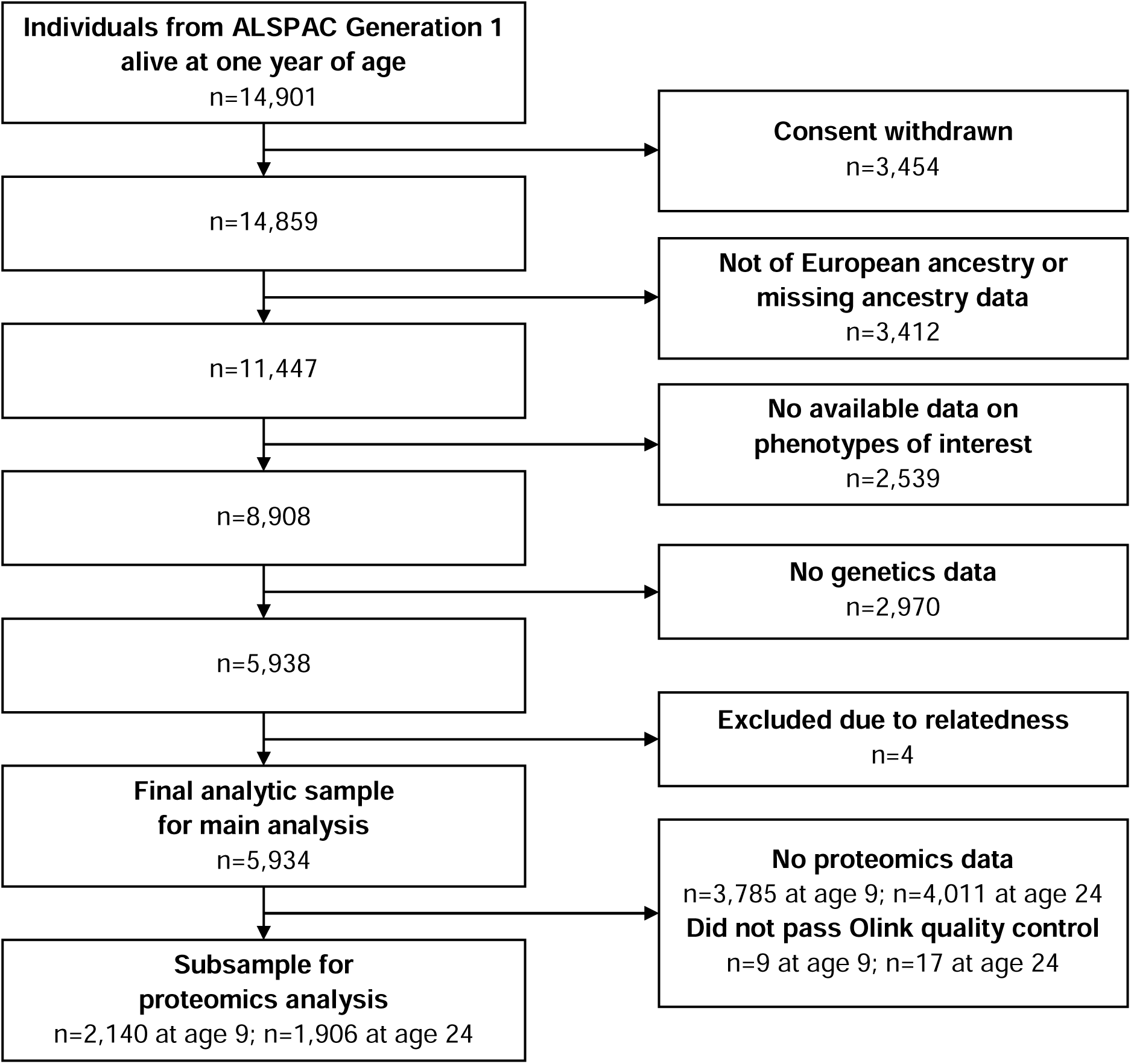
Flow diagram of study sample selection from the Avon Longitudinal Study of Parents and Children (ALSPAC).

**Table 1.** Sociodemographic and health characteristics of included individuals from the Avon Longitudinal Study of Parents and Children (ALSPAC).

|  | Available n | n (%) or median [IQR] |
|---|---|---|
| Sex at birth: Female | 5934 | 2978 (50.2%) |
| Maternal education | 5919 |  |
| CSE |  | 779 (13.2%) |
| Vocational |  | 497 (8.4%) |
| O-level |  | 2053 (34.7%) |
| A-level |  | 1569 (26.5%) |
| Degree |  | 1021 (17.2%) |
| AUDIT-C at age 24 | 2440 | 5 [4, 7] |
| Smoker at age 24 | 2466 | 700 (28.4%) |
| Depressive episode at age 24 | 2464 | 253 (10.3%) |
| Generalized anxiety disorder at age 24 | 2460 | 241 (9.8%) |

Depressive episode and generalized anxiety disorder were assessed using the Clinical Interview Schedule – Revised (CIS-R).

AUDIT-C: Alcohol Use Disorders Identification Test for Consumption. A score of 5 or more on the AUDIT-C is a positive screen, indicating drinking levels that are potentially harmful for health.

### Main longitudinal analysis

We observed ICM-MM PRS main effects, with a higher ICM-MM PRS associated with higher BMI, FMI, LMI, SBP, DBP, FI and CRP, but lower HDL at their respective baselines. This corresponded to a 1.48% (95% CI 1.11%-1.86%) higher BMI, 1.31% (95% CI 1.11%-1.52%) higher SBP and 1.00% (95% CI 0.74%-1.27%) higher DBP at age 7; 7.02% (95% CI 5.59%-8.46%) higher FMI, 0.29% (95% CI 0.06%-0.52%) higher LMI, 9.81% (95% CI 5.84%-13.93%) higher CRP and 0.89% (95% CI 0.28%-1.50%) lower HDL at age 9; and 3.17% (95% CI 1.12%-5.21%) higher FI at age 15 with each standard deviation increase in ICM-MM PRS. There were also age x PRS interaction effects on mental and cardiometabolic outcomes, suggesting that a higher ICM-MM PRS was associated with faster increases in depressive symptoms, BMI, LMI, non-HDL, TG and FI, faster decreases in SBP and slower increases in HDL. Details on model selection and final models are described in **Tables S5 and S6**, and results are presented in **Figure 2** and **Table 2**. We observed sex differences across all phenotypes of interest, with females having higher depressive symptoms, BMI, FMI, DBP, HDL, non-HDL, TG, FI and CRP, and lower LMI, SBP and FG at baseline.

**Figure 2.**
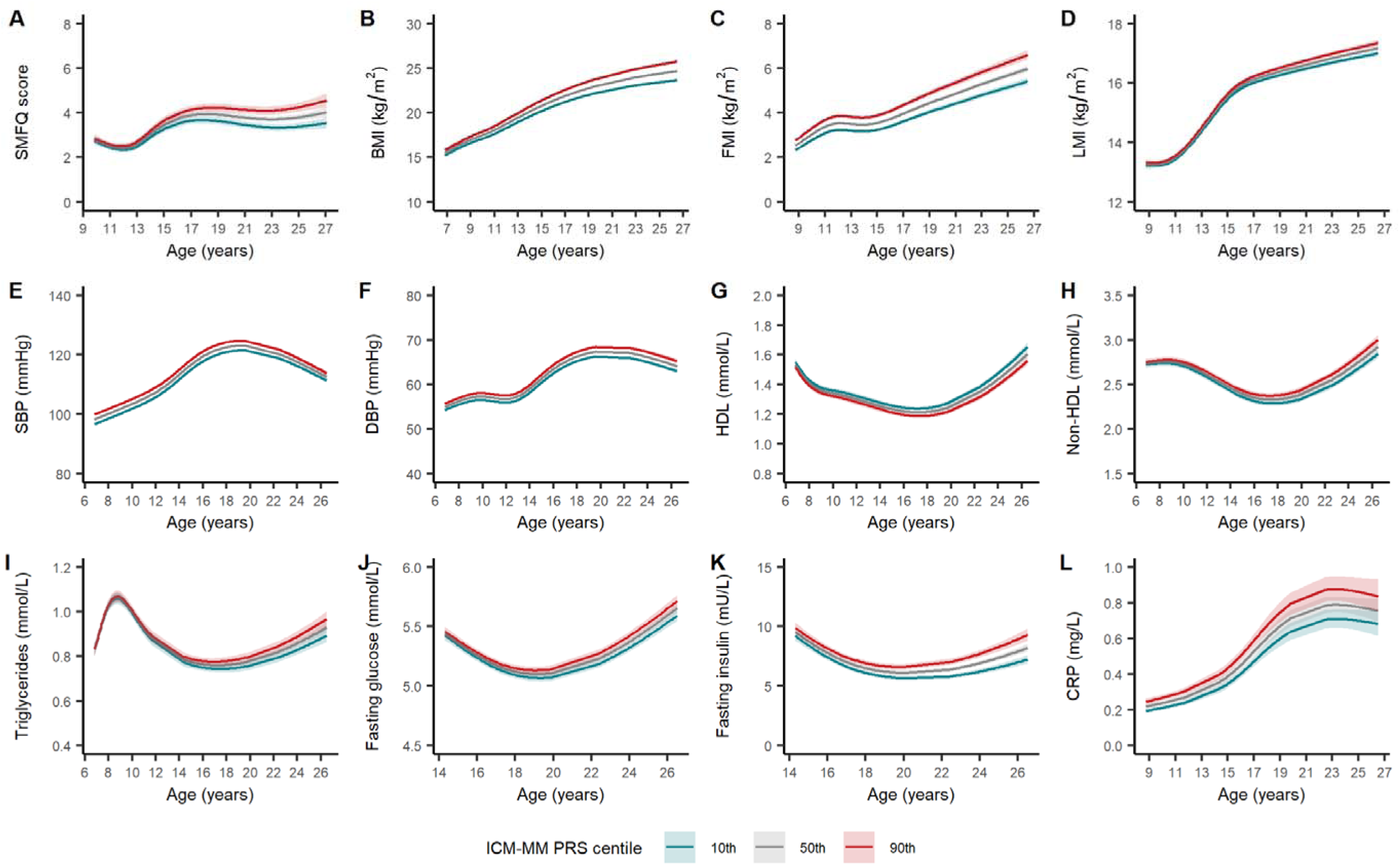
Predicted mental and cardiometabolic health trajectories for individuals at the 10^th^, 50^th^ and 90^th^ percentile of the adult internalizing-cardiometabolic multimorbidity polygenic risk score (ICM-MM PRS) in the Avon Longitudinal Study of Parents and Children (ALSPAC). Panel A – depressive symptoms; B – body mass index; C – fat mass index; D – lean mass index; E – systolic blood pressure; F – diastolic blood pressure; G – high-density lipoprotein; H – non-high-density lipoprotein; I – triglycerides; J – fasting glucose; K – fasting insulin; L – C-reactive protein.

**Table 2.** Main and interaction effects of the adult internalizing-cardiometabolic multimorbidity polygenic risk score (ICM-MM PRS) on mental and cardiometabolic health outcomes.

| Outcome | Term | Estimate | Lower CI | Upper CI | *p*-value | Adjusted *p*-value |
|---|---|---|---|---|---|---|
| Depressive symptoms | PRS | 1.0122 | 0.9934 | 1.0313 | 2.04E-01 | 2.33E-01 |
|  | **Age x PRS** | **1.0039** | **1.0024** | **1.0055** | **8.77E-07** | **3.01E-06** |
| BMI | **PRS** | **1.0148** | **1.0111** | **1.0186** | **8.91E-15** | **5.35E-14** |
|  | **Age x PRS** | **1.0010** | **1.0008** | **1.0011** | **1.91E-32** | **2.29E-31** |
| FMI | **PRS** | **1.0702** | **1.0559** | **1.0846** | **4.90E-23** | **3.92E-22** |
|  | Age x PRS | 1.0006 | 0.9997 | 1.0014 | 1.74E-01 | 2.09E-01 |
| LMI | **PRS** | **1.0029** | **1.0006** | **1.0052** | **1.32E-02** | **2.12E-02** |
|  | **Age x PRS** | **1.0003** | **1.0001** | **1.0005** | **1.72E-03** | **3.98E-03** |
| Systolic blood pressure | **PRS** | **1.0131** | **1.0110** | **1.0152** | **1.30E-34** | **3.13E-33** |
|  | **Age x PRS** | **0.9998** | **0.9996** | **1.0000** | **2.24E-02** | **3.37E-02** |
| Diastolic blood pressure | **PRS** | **1.0100** | **1.0074** | **1.0127** | **1.11E-13** | **5.32E-13** |
|  | Age x PRS | 1.0002 | 0.9999 | 1.0005 | 1.34E-01 | 1.70E-01 |
| HDL | **PRS** | **0.9911** | **0.9850** | **0.9972** | **4.64E-03** | **7.95E-03** |
|  | **Age x PRS** | **0.9993** | **0.9988** | **0.9998** | **3.69E-03** | **6.81E-03** |
| Non-HDL | PRS | 1.0029 | 0.9981 | 1.0078 | 2.35E-01 | 2.56E-01 |
|  | **Age x PRS** | **1.0007** | **1.0003** | **1.0010** | **3.66E-04** | **9.76E-04** |
| Triglycerides | PRS | 1.0006 | 0.9895 | 1.0118 | 9.15E-01 | 9.15E-01 |
|  | **Age x PRS** | **1.0016** | **1.0005** | **1.0026** | **3.51E-03** | **6.81E-03** |
| Fasting glucose | PRS | 1.0029 | 0.9997 | 1.0061 | 7.69E-02 | 1.03E-01 |
|  | Age x PRS | 1.0005 | 1.0000 | 1.0010 | 4.16E-02 | 5.87E-02 |
| Fasting insulin | **PRS** | **1.0317** | **1.0117** | **1.0521** | **1.82E-03** | **3.98E-03** |
|  | **Age x PRS** | **1.0059** | **1.0029** | **1.0090** | **1.39E-04** | **4.16E-04** |
| CRP | **PRS** | **1.0981** | **1.0584** | **1.1393** | **6.36E-07** | **2.54E-06** |
|  | Age x PRS | 0.9993 | 0.9954 | 1.0031 | 7.04E-01 | 7.35E-01 |

Estimates reflect percentage change per one standard deviation increase in ICM-MM PRS.
Results in bold indicate associations with Benjamini-Hochberg adjusted *p*-values < 0.05.
BMI – body mass index; CRP – C-reactive protein; FMI – fat mass index; HDL – high-density lipoprotein; LMI – lean mass index.

### Sensitivity analysis

Results from the sensitivity analyses using the ICM-MM PRS_GPLC_, all component single-trait PRSs and a negative control PRS suggest our findings are stable. Overall patterns of associations in the sensitivity analyses involving the ICM-MM PRS_GPLC_ (**Table S7**) and BMI PRS (**Table S9**) were similar to those in the main analysis, though none of the PRSs examined consistently showed stronger associations with all traits of interest than others.

Other component single-trait PRSs showed fewer associations with all traits of interest (**Tables S8, S10-S14)**. The negative control (age-related macular degeneration PRS_GPLC_) did not show evidence of strong association with outcomes of interest (**Table S15**). While the effect size decreased, the association between the ICM-MM PRS and CRP remained after excluding CRP values >10mg/L (**Table S16**).

### Secondary analysis

The ICM-MM PRS was associated with higher levels of 18 proteins at age 9, and higher levels of 10 proteins and lower levels of 2 proteins at age 24. Associations that were consistent across both time points are higher levels of vascular endothelial growth factor A (VEGFA), monocyte chemotactic protein 3 (MCP-3 [also known as C-C motif chemokine ligand 7 or CCL7]), interleukin-6 (IL-6), tumor necrosis factor superfamily member 14 (TNFSF14), fibroblast growth factor 21 (FGF-21), interleukin-18 receptor 1 (IL18R1), and hepatocyte growth factor (HGF). Associations with IL-6, TNFSF14 and HGF remained after correction for multiple comparisons (**Figure 3** and **Table S17**). Sensitivity analyses with component single-trait PRSs revealed only a consistent association between the BMI PRS and IL-6.

**Figure 3.**
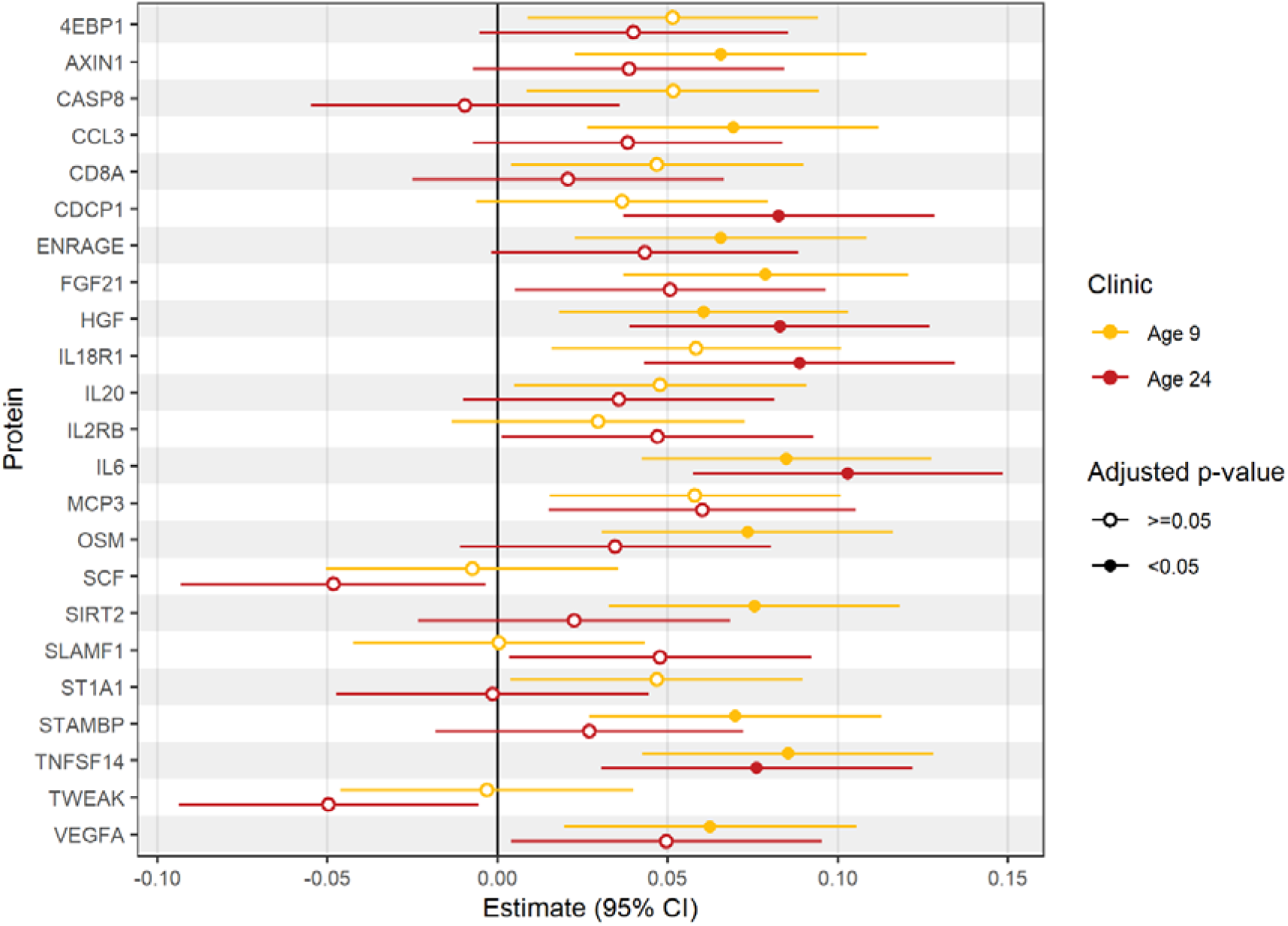
Dot-and-whisker plots for associations of the adult internalizing-cardiometabolic multimorbidity polygenic risk score (ICM-MM PRS) with circulating immune proteins at ages 9 and 24. 4EBP1 – eukaryotic translation initiation factor 4E-binding protein 1; AXIN1 – Axin-1; CASP8 – caspase-8; CCL3 – C-C motif chemokine 3; CD8A – T-cell surface glycoprotein CD8 alpha chain; CDCP1 – CUB domain-containing protein 1; ENRAGE – protein S100-A12; FGF21 – fibroblast growth factor 21; HGF – hepatocyte growth factor; IL18R1 – interleukin-18 receptor 1; IL20 – interleukin-20; IL2RB – interleukin-2 receptor subunit beta; IL6 - interleukin-6; MCP3 – monocyte chemotactic protein 3; OSM – oncostatin-M; SCF – stem cell factor; SIRT2 – SIR2-like protein 2; SLAMF1 – signalling lymphocytic activation molecule; ST1A1 – sulfotransferase 1A1; STAMBP – STAM-binding protein; TNFSF14 – tumour necrosis factor ligand superfamily member 14; TWEAK – tumour necrosis factor ligand superfamily member 12; VEGFA – vascular endothelial growth factor A.

The protein-protein interaction network suggested substantial interactions and most proteins fell into one main cluster associated with IL-6 (**Figure S2**). Pathway analysis showed that these proteins are primarily involved in immune cell chemotaxis and migration.

## Discussion

### Key findings

In this study, we examined the effects of polygenic risk of adult ICM-MM on longitudinal changes in 12 mental and cardiometabolic health outcomes from childhood to early adulthood. We observed that a higher ICM-MM PRS was associated with higher BMI, FMI, LMI, SBP, DBP, FI and CRP, with the largest effects observed for CRP and FMI. Moreover, higher ICM-MM PRS was associated with faster increases in depressive symptoms, BMI, LMI, non-HDL, TG and FI, faster decreases in SBP and slower increases in HDL during certain periods of development, generally between puberty and early adulthood. These findings suggest that differences in adiposity and adiposity-driven inflammation could be some of the earliest manifestations of ICM-MM polygenic liability in a population-based sample, with smaller effects on some other cardiometabolic traits emerging in adolescence and early adulthood. The pattern of associations in models with the BMI PRS was similar, suggesting these effects may be driven by genetic influences on BMI.

Given the effect on CRP, an archetypal systemic inflammatory marker, we further examined associations of the ICM-MM PRS with a range of circulating immune response-related proteins in childhood and early adulthood. We identified higher levels of three proteins (IL-6, TNFSF14 and HGF) as associated with higher polygenic liability of ICM-MM at both time points. These proteins are involved in cytokine signaling, cell chemotaxis, migration and chemokine production.

### Comparison to existing literature

To our knowledge, this is the first study to investigate the effects of polygenic liability of a specific type of adult multimorbidity on early-life health trajectories. Cohort studies with rich longitudinal phenotypic data are scarce, so there are few studies examining long-term changes in multiple health outcomes using a life course epidemiological approach. The shapes of the trajectories characterized here are consistent with earlier work conducted using ALSPAC data (45–47).

A large proportion of the limited literature on polygenic liability of disease and health trajectories is focused on psychiatric disorders. Several earlier studies have shown that major depressive disorder PRS is associated with specific depression trajectories, particularly an early-onset persistent depression trajectory (48–51). In contrast, studies examining associations between genetic risk and cardiometabolic risk factor trajectories report mixed findings, with some describing associations only at specific time points or with mean levels but not change over time (e.g., blood pressure PRS on SBP (52) and Alzheimer’s disease PRS on cardiometabolic risk factors (53)), whereas others also showed associations with change over time (e.g., BMI PRS on growth parameters (54), BMI PRS on weight (28), and body size PRS on DXA-derived fat and lean mass (55)).

Research on biomarkers of multimorbidity is sparse, and there are no studies that specifically focus on ICM-MM. Earlier systematic reviews reported that various types of biomarkers have been investigated in relation to multimorbidity (general or specific clusters), but most have been studied in one or two studies only (56, 57). A recent updated systematic review suggested that a range of routinely assessed biomarkers of inflammation, metabolism and neurodegeneration may help detect or predict multimorbidity (58). Specifically, the review identified IL-6, glycated hemoglobin (HbA1c) and neurofibrillary tangles as key biomarkers for multimorbidity. Associations with some other biomarkers (e.g., lipids and markers of kidney and liver function) have also been found, but findings were inconsistent.

The systematic review recommended that future studies investigate progression trends using longitudinal designs and try to disentangle biological interactions using multi-omics data. One recent study that used high-throughput proteomics data examined associations of plasma proteins with incident multimorbidity in UK Biobank, and identified 233 proteins associated with incident multimorbidity, which included IL-6 and HGF (59).

The proteins identified in our study (IL-6, TNFSF14 and HGF) have previously been implicated (including evidence for potential causal roles) in some of the individual conditions that are part of our ICM-MM definition. IL-6 is a pleiotropic immunomodulatory cytokine that has roles in the regulation of inflammation and can have either pro-or anti-inflammatory effects via classical, trans-or cluster signaling. Circulating IL-6 has been shown to be potentially causally linked to depression (60, 61) and type 2 diabetes (62), but not anxiety (61, 63). Findings on the causal effects of IL-6 on blood pressure traits were mixed (64).

TNFSF14 is an immunostimulatory cytokine that can either bind to the herpes simplex virus mediator (HVEM) to amplify immune responses or bind to the lymphotoxin-β receptor to induce apoptosis. Preclinical research suggests that TNFSF14 modulates adipose tissue inflammatory responses through enhancing macrophage/T cell infiltration and activation (65), and can induce hypertension in both pregnant and non-pregnant mice (66). Upregulation of TNFSF14 was observed in major depressive disorder (67, 68) and type 2 diabetes (69). It has been postulated that TNFSF14 plays regulatory roles in glucose homeostasis and lipid metabolism (70, 71), and therefore may be a novel target for obesity and cardiometabolic disease. However, potential causal associations of TNFSF14 with internalizing or cardiometabolic disease have not been investigated.

HGF is an adipokine that binds exclusively to the MET receptor and is involved in a range of biological processes including cell motility, survival and proliferation. Observational studies have reported associations of HGF with major depressive disorder (67, 68), various cardiometabolic disease (72, 73) and insulin resistance (74), with Mendelian randomization studies suggesting a causal role for circulating HGF in the development of coronary heart disease (75) as well as LDL cholesterol, total cholesterol and triglycerides (76).

There is now a large body of evidence supporting immune-mediated mechanisms of atherosclerosis (77–79). Given the roles of these identified proteins in cytokine signaling, cell chemotaxis and migration and chemokine production, they may underlie plaque development and progression in atherosclerosis, and pathogenesis of subsequent cardiometabolic disease and multimorbidity.

Lastly, there is a large body of literature supporting a bidirectional relationship between obesity and depression (80, 81), with inflammation or immunometabolic dysregulation potentially mediating this relationship (82, 83). The relationship between depression and inflammation is also likely bidirectional (84), and both BMI and depression may drive increases in inflammatory markers, including the immune proteins identified in this study (85–88). Taken together, it suggests targeting overweight/obesity and inflammation in young people may be promising avenues to reduce or prevent future ICM-MM.

### Implications

Internalizing and cardiometabolic conditions collectively account for a large proportion of health-related morbidity and mortality globally. Our findings suggest that adiposity and adiposity-related inflammation could be some of the earliest manifestations of ICM-MM polygenic liability in young people. These results support the potential of obesity and inflammation as targets for prevention and intervention efforts to reduce the risk of adult multimorbidity, particularly for internalizing and cardiometabolic conditions.

### Limitations

There are some limitations to this study. Most of the discovery GWASs used for the calculation of the ICM-MM PRS were conducted in European cohorts, and the sample of this present study is also restricted to individuals reported by their parents to be of European ancestry, which limits generalizability of our findings to other populations. As with all longitudinal cohorts, there is attrition in ALSPAC, which previous research has shown to not be at random and dropout is systematically related to various family adversity variables (89); however, bias due to missing outcome data may have been reduced by including all outcome data from all eligible individuals in the trajectory models. The models used in this study assume there is only one overall trajectory in the population and individuals deviate from this only in their slopes and intercepts; however, this may not necessarily be true across all phenotypes of interest (e.g., there are latent depressive symptom trajectories of different shapes (90)). While there is evidence to suggest that there are sex differences in cardiometabolic health trajectories in development (45), the current sample size is likely underpowered to detect polygenic effects in a stratified analysis. Lastly, there is a possibility that genetic contributions to the health outcomes investigated vary with age, as has been shown with BMI (91), and the specific genetic variants influencing these outcomes and their effects may differ across different ages (92), which have not been accounted for in our present analysis.

## Conclusions

In this study, we leveraged rich longitudinal data from a large birth cohort to examine early manifestations of ICM-MM polygenic liability. We identified tangible life course signals, with the strongest effects observed in anthropometric, body composition and inflammatory measures, highlighting potential targets for multimorbidity risk reduction or prevention.

Furthermore, we identified three proteomic markers linked to ICM-MM polygenic risk, which provides some insights into potential biological mechanisms and pathways that underlie ICM-MM. Our work provides a framework to better understand how and when risk for multimorbidity develops from childhood through to adulthood, which can be adapted to study other risk factors or other types of multimorbidity.

## Acknowledgements

We are extremely grateful to all the families who took part in this study, the midwives for their help in recruiting them, and the whole ALSPAC team, which includes data collection staff, data and administration staff, technical managers and the technical staff with the Bristol Bioresource Laboratory, based within the University of Bristol. We would also like to thank members of the LINC study public advisory group for their valuable contribution.

This work was funded by the Tackling Multimorbidity at Scale Strategic Priorities Fund programme [grant number MR/W014416/1] delivered by the Medical Research Council and the National Institute for Health Research in partnership with the Economic and Social Research Council and in collaboration with the Engineering and Physical Sciences Research Council. RSMT, DS and IKK were supported by this grant.

GMK acknowledges funding support from the UK Medical Research Council (MRC), grant number: MC_UU_00032/6, which forms part of the MRC Integrative Epidemiology Unit at the University of Bristol. This grants supports RT. GMK also acknowledges funding from the Wellcome Trust (grant numbers: 201486/Z/16/Z and 201486/B/16/Z), the Medical Research Council (grant numbers: MR/W014416/1; MR/S037675/1; MR/Z50354X/1; and MR/Z503745/1), and the UK National Institute of Health and Care Research (NIHR) Bristol Biomedical Research Centre (grant number: NIHR 203315). MBMvdB acknowledges funding from the UK Medical Research Council (MRC)(MR/W028395/1; MR/W020297/1; MR/V004905/1RE; MR/T033045/1; MR/S037667/1), Wellcome Trust (W227882/Z/23/Z and 226709/Z/22/Z) and The National Institutes of Mental Health (U01MH119758).

The views expressed are those of the authors and not necessarily those of the UK NIHR or the Department of Health and Social Care.

The UK Medical Research Council and Wellcome (Ref: MR/Z505924/1) and the University of Bristol provide core support for ALSPAC. This publication is the work of the authors and RSMT, GMK and NJT will serve as guarantors for the contents of this paper. A comprehensive lists of grants funding is available on the ALSPAC website (https://www.bristol.ac.uk/alspac/external/documents/grant-acknowledgements.pdf).

Genome-wide genotyping data were generated by Sample Logistics and Genotyping Facilities at Wellcome Sanger Institute and LabCorp (Laboratory Corporation of America) using support from 23andMe.

## LINC Consortium members

Marianne B. M. van den Bree, George Kirov, Jack F. G. Underwood, Michael J. Owen, James T. R. Walters, Peter A. Holmans, Jane Lynch, Ioanna K. Katzourou (Cardiff University, UK)

David A. van Heel, Sarah Finer, Daniel Stow (Queen Mary University of London, UK)

Golam M. Khandaker, Nicholas J. Timpson, John A. A. Macleod, Julie P. Clayton, Ruby S. M. Tsang, Jane Sprackman, Shahid Khan (University of Bristol, UK)

Inês Barroso, Rupert A. Payne (University of Exeter, UK) Mark Mon-Williams, Nabila Ali (University of Leeds, UK) Hilary C. Martin (Wellcome Sanger Institute, UK)

Thomas Werge, Andrés Ingason, Morteza Vaez, Lam O. Huang (Institute of Biological Psychiatry, Denmark)

## Data availability statement

The informed consent obtained from ALSPAC (Avon Longitudinal Study of Parents and Children) participants does not allow the data to be made available through any third party maintained public repository. Supporting data are available from ALSPAC on request under the approved proposal number, B4721. Full instructions for applying for data access can be found here: https://www.bristol.ac.uk/alspac/researchers/access/. The ALSPAC study website contains details of all available data (https://www.bristol.ac.uk/alspac/researchers/our-data/).

## Code availability

R scripts used for the analyses can be found at: https://github.com/rubietsang/multimorbidity-prs-health-trajectories

