## Supplementary Material for "Early-life hallmarks of polygenic liability to adult internalizing-cardiometabolic multimorbidity"

**Supplementary Methods**

Additional information on data collection

Study data were collected and managed using REDCap electronic data capture tools hosted at the University of Bristol (1). REDCap (Research Electronic Data Capture) is a secure, web-based software platform designed to support data capture for research studies.

Additional information on outcome variables

*Alcohol use and smoking*

Alcohol use was assessed using the Alcohol Use Disorders Identification Test for Consumption (AUDIT-C) (2) at the age 24 clinic. This consists of three questions on frequency and amount of alcohol consumption, and each item are scored on a scale of zero to four. A score of five or above is a positive screen for risky alcohol use. A composite smoking variable was derived by coding those who self-reported having smoked a cigarette in the past 30 days and at least one cigarette per day or at least seven cigarettes per week as smokers.

*Depressive episode and generalised anxiety disorder*

ICD-10 depressive episode and generalised anxiety disorder were derived from the self-administered Clinical Interview Schedule – Revised (CIS-R) (3). The interview includes general questions to establish an overall picture of health, followed by 14 sections each scoring for a particular type of neurotic symptom, namely somatic symptoms, fatigue, concentration and forgetfulness, sleep problems, irritability, worry about physical health, depression, depressive ideas, worry, anxiety, phobias, panic, compulsions, and obsessions. An algorithm is then applied to derive a number of ICD-10 diagnoses for neuropsychiatric disorders.

*Depressive symptoms*

Depressive symptoms were assessed using the self-reported 13-item Short Mood and Feelings Questionnaire (SMFQ) (4) (range 0-26, higher scores indicate more depressive symptoms). Questions were answered based on the two weeks prior to completing the questionnaire. For individuals with missing data on fewer than three questions, the total was computed with median replacement for the missing items. For each occasion, individuals with missing data on more than three questions had their total score recoded as missing. We included data collected from 10 occasions between the ages of 10 and 25 years (approximately at ages 10, 12, 13, 16, 17, 18, 21, 22, 23, 25), ending with the last questionnaire administered in 2017-2018 prior to the start of the COVID-19 pandemic.

*Anthropometric and body composition measures*

Height and weight were measured at all clinic visits. Height was measured to the nearest 0.1cm using a Harpenden stadiometer (Holtain Ltd., Crymych, Pembrokeshire, UK). Weight was measured to the nearest 0.1kg using Tanita scales (Tanita UK Ltd., Yewsley, Middlesex, UK). A whole body dual-energy X-ray absorptiometry (DXA) scan was performed at the age 9, 11, 13, 15, 17 and 24 clinics using a Lunar Prodigy narrow fan beam densitometer (GE Medical System Lunar, Madison, WI, USA), where bone content, lean and fat masses were measured. Body mass index (BMI), total body lean mass index and fat mass index were calculated as weight (kg), total body lean mass and total body fat mass divided by height squared (m^2^) respectively.

*Blood pressure*

Blood pressure was measured at the age 7, 9, 11, 12, 13, 15, 17 and 24 clinics using an automatic oscillometric device (Dinamap 9301 Vital Signs Monitor at ages 7, 9, 11, 13, 15 and 17, and a Dinamap 8100 Vital Signs Monitor at age 12). At each clinic, systolic and diastolic blood pressure were measured twice each, from the right arm where possible, with the participant sitting and at rest with the arm supported, using a cuff size appropriate for the participant’s upper arm circumference. A mean was calculated from the two readings.

*Lipids*

Non-fasting blood samples were collected at ages 7 and 9 years and fasting blood samples at ages 15, 17 and 24 years, and immediately spun and frozen at -80°C. Total cholesterol, triglycerides and high-density lipoprotein cholesterol (HDL) were assayed by modification of the standard Lipid Research Clinics Protocol using enzymatic reagents for lipid determination. Non-HDL was calculated by subtracting HDL from total cholesterol.

*Immunometabolic biomarkers*

Fasting glucose was measured by hexokinase method at ages 15, 17 and 24. Fasting insulin was measured using an enzyme linked immunosorbent assay (ELISA) kit (Mercodia, Uppsala, Sweden) at ages 15, 17, and an electrochemiluminescence immunoassay (ECLIA) kit (Roche Diagnostics, Mannheim, Germany) at age 24. C-reactive protein (CRP) was measured by automated particle-enhanced immunoturbidimetric assay (Roche Diagnostics Corporation, Indianapolis, IN, USA) at ages 9, 15, 17 and 24.

Additional information on informed consent

The completion of a questionnaire, either on paper or online, was considered to be written consent from participants to use their data for research purposes. For the majority of tests undertaken during face-to-face visits, verbal consent was obtained from participants (both parents and children as appropriate) prior to the start of any data collection. However, some tests required the completion of a written consent form. Biological samples are collected in accordance with the Human Tissue Act (2004). Specific Research Ethics Committee approval is sought for the consenting process at each collection sweep. Written consent, including permission for future use, is obtained from adult participants or from the parents of children as appropriate. Ethical approval for future use is covered by ALSPAC’s Research Tissue Bank approval. All historical consents to hold biological samples have been reviewed as part of the Tissue Bank approval process. Participants can contact the study team at any time to retrospectively withdraw consent for use of their samples.

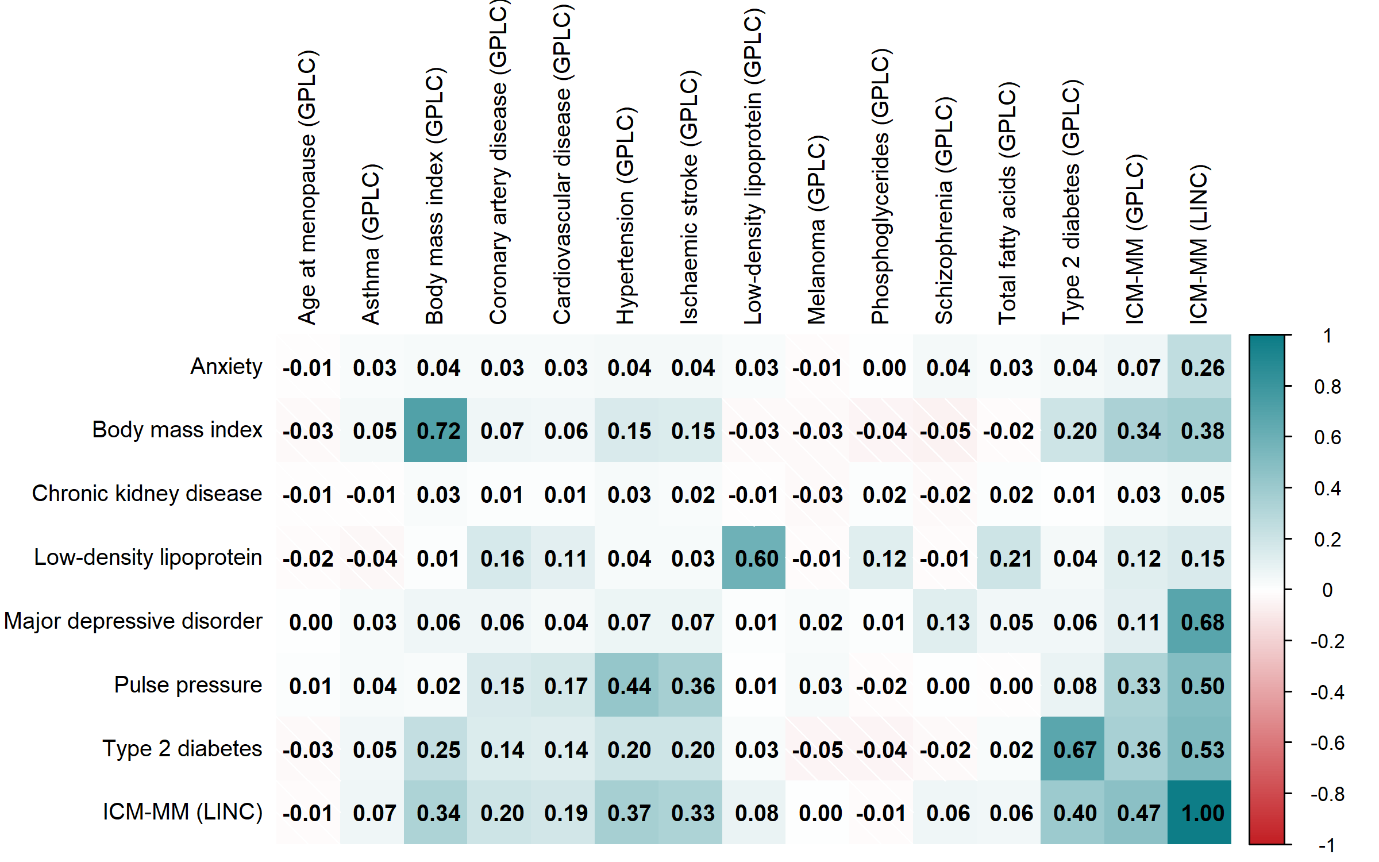

**Figure S1. Heatmap of Pearson correlations between ICM-MM PRS and ICM-MM PRS_GPLC_ and their components in ALSPAC.**

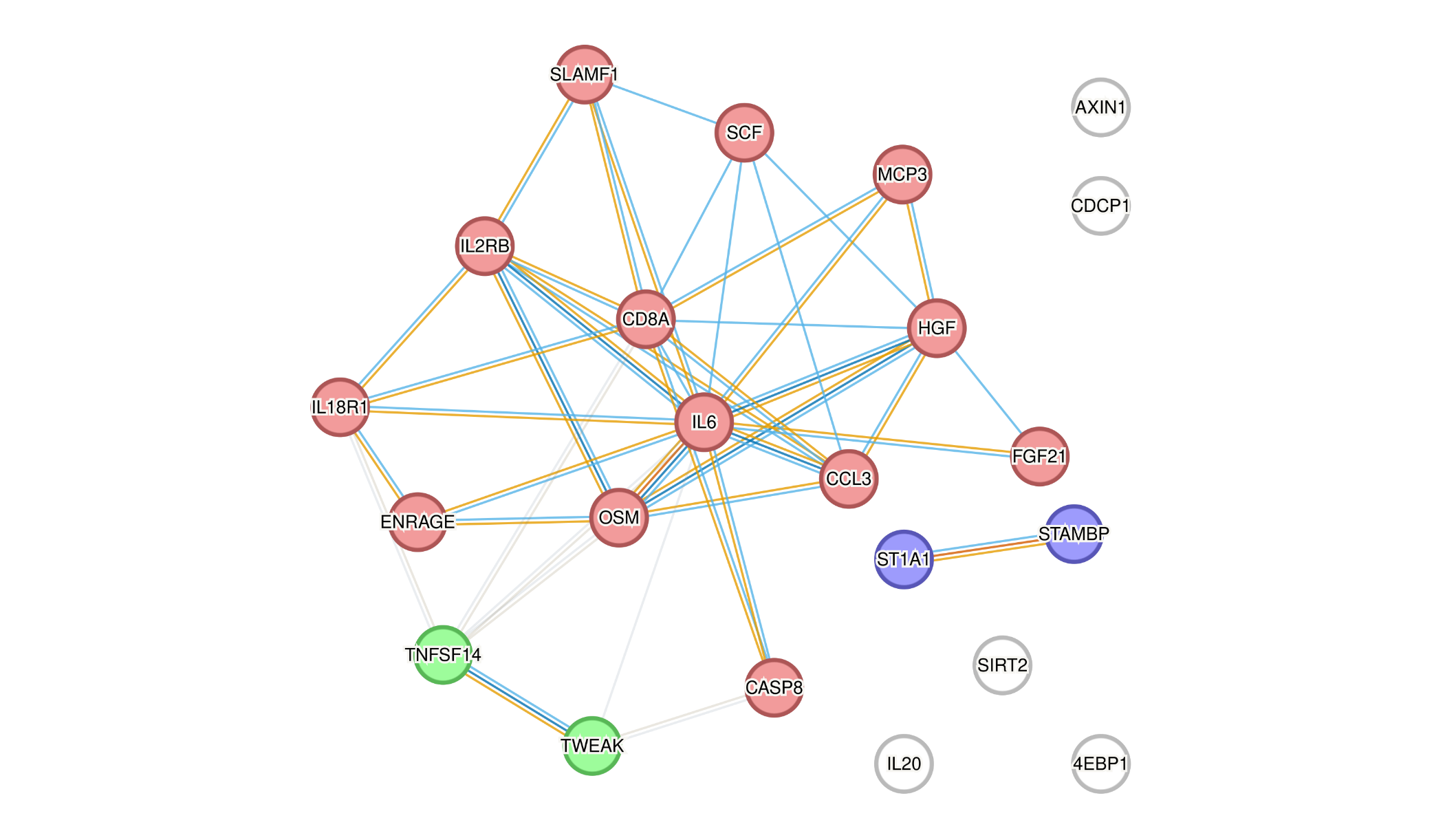

**Figure S2. Protein-protein interaction network of the identified immune and inflammatory proteins associated with ICM-MM polygenic risk (parameters used: full STRING network, network edges for evidence, including all active interaction sources, with a minimum required interaction score of 0.400; clustering was performed using k-means clustering for 3 clusters).** 4EBP1 – eukaryotic translation initiation factor 4E-binding protein 1; AXIN1 – Axin-1; CASP8 – caspase-8; CCL3 – C-C motif chemokine 3; CD8A – T-cell surface glycoprotein CD8 alpha chain; CDCP1 – CUB domain-containing protein 1; ENRAGE – protein S100-A12; FGF21 – fibroblast growth factor 21; HGF – hepatocyte growth factor; IL18R1 – interleukin-18 receptor 1; IL20 – interleukin-20; IL2RB – interleukin-2 receptor subunit beta; IL6 - interleukin-6; MCP3 – monocyte chemotactic protein 3; OSM – oncostatin-M; SCF – stem cell factor; SIRT2 – SIR2-like protein 2; SLAMF1 – signalling lymphocytic activation molecule; ST1A1 – sulfotransferase 1A1; STAMBP – STAM-binding protein; TNFSF14 – tumour necrosis factor ligand superfamily member 14; TWEAK – tumour necrosis factor ligand superfamily member 12. Note: VEGFA – vascular endothelial growth factor A was not included in version 12.5.
 **References**

1.	Harris PA, Taylor R, Thielke R, Payne J, Gonzalez N, Conde JG (2009): Research electronic data capture (REDCap)—A metadata-driven methodology and workflow process for providing translational research informatics support. *Journal of Biomedical Informatics*. 42:377-381.

2.	Bush K, Kivlahan DR, McDonell MB, Fihn SD, Bradley KA, for the Ambulatory Care Quality Improvement P (1998): The AUDIT Alcohol Consumption Questions (AUDIT-C): An Effective Brief Screening Test for Problem Drinking. *Archives of Internal Medicine*. 158:1789-1795.

3.	Lewis G, Pelosi AJ, Araya R, Dunn G (1992): Measuring psychiatric disorder in the community: a standardized assessment for use by lay interviewers. *Psychological Medicine*. 22:465-486.

4.	Angold A, Costello EJ, Messer SC, Pickles A (1995): Development of a short questionnaire for use in epidemiological studies of depression in children and adolescents. *International Journal of Methods in Psychiatric Research*. 5:237-249.
